# Sex Differences in Endocannabinoid and Inflammatory Markers Associated with Posttraumatic Stress Disorder

**DOI:** 10.1101/2025.01.13.25320467

**Authors:** Therese A. Rajasekera, Anna Joseph, Hui Pan, Jonathan M. Dreyfuss, Doruntina Fida, Julie Wilson, Madeline Behee, Raina N Fichorova, Resat Cinar, Primavera A. Spagnolo

## Abstract

**Background:** Posttraumatic stress disorder (PTSD) is one of the most sex-polarized psychiatric disorders, with women exhibiting twice the prevalence of men. The biological mechanisms underlying this sex disparity are not fully understood. Growing evidence suggests that alterations of the stress-buffering endocannabinoid (eCB) system and heightened inflammation are central to PTSD pathophysiology and may contribute to sex-biases in PTSD risk and severity. Here, we examined sex-differences in levels of circulating eCBs and peripheral pro-inflammatory markers in a cohort of men and women with PTSD and non-psychiatric controls.

**Methods:** 88 individuals with PTSD and 85 sex- and age-matched controls (HC) were retrospectively selected from the Mass General Brigham Biobank. Serum was assayed to measure circulating eCBs [anandamide (AEA), 2-arachidonyl glycerol (2-AG), oleoylethanolamide (OEA), and arachidonic acid (AA] and inflammatory markers [interleukin-1β (IL-1β), IL-6, IL-8, IL-18, tumor necrosis factor-alpha (TNFα), and C-reactive Protein (CRP)]. Linear regression was used to predict differential abundance of each analyte by disease state (PTSD/HC) within the male and female subgroups. Two-sided t-tests with Benjamini-Hochberg correction were used to examine differences across subgroups. Analyses were repeated in those with comorbid major depressive disorder.

**Results:** Male PTSD patients showed a significant decrease in AEA, AA and OEA levels compared to male controls (*p*’s < .001) and to female subgroups (PTSD and HCs) (*p< .001*). In contrast, among females, PTSD patients showed elevated levels of IL-6 (*p<.05*) and IL-8 (*p<.05*). Both male and female PTSD patients exhibited an increase in TNFα concentrations (*p<.05*), compared to HCs. Similar results were obtained in the subgroup of individuals with comorbid MDD and after controlling for the *FAAH* 385A genotype.

**Conclusions:** Our findings show for the first time that decrease in eCB levels is absent in women with PTSD, who in turn exhibit a broader increase in inflammatory markers, thus suggesting that biological perturbations underlying PTSD may vary by sex.

## 1. Introduction

Posttraumatic stress disorder (PTSD) is a maladaptive and debilitating psychiatric disorder that can develop following exposure to severe trauma. In the United States, PTSD affects 5-10% of adults, with women exhibiting twice the prevalence of men^1–3^. Furthermore, women with PTSD experience more severe symptoms and a higher risk of comorbid conditions, including major depressive disorder, cardiometabolic diseases, and autoimmune disorders^4^. Despite these well-documented disparities, the biological mechanisms underlying sex differences in PTSD risk and presentation remain poorly understood.

Sex hormones, particularly estradiol, have traditionally been viewed as primary contributors to these sex differences. Evidence from both animal and human studies have demonstrated that physiological fluctuations in estradiol levels exert a profound and dynamic modulation on the hypothalamic-pituitary-adrenal (HPA) axis and on the neural machinery of fear processing, contributing to an increased risk for PTSD in females^5^. However, recent research suggests that other biological mechanisms may be implicated in fear and stress dysregulations observed in PTSD and that such mechanisms may contribute to PTSD pathophysiology in a sex-specific manner. Specifically, perturbations in the endocannabinoid (eCB) system and heightened inflammation have been consistently reported in individuals affected by PTSD^6–9^.

For example, circulating levels of anandamide, one of the main eCB ligands, are significantly reduced in individuals with PTSD compared to both healthy controls and those exposed to trauma that did not develop PTSD^10^. Conversely, increased anandamide (AEA) levels have been associated with decreased anxiety in response to a stress-provoking experimental procedure and with reduced severity of hyperarousal symptoms in a sample of subjects with comorbid PTSD and alcohol use disorder^11^. In addition, pharmacological inhibition of fatty-acid amide hydrolase (FAAH), the primary enzyme capable of AEA hydrolysis, improves the recall of fear extinction memories and attenuates the anxiogenic effects of stress in a sample of healthy individuals^12^.

However, recent preclinical studies have provided evidence that AEA signaling differentially affects fear expression and extinction in males and females, highlighting potential sex-based differences in the role of the eCB system in regulating fear and stress responses^13^. These findings are supported by human research showing that AEA and other endocannabinoid levels vary between men and women across the lifespan^14^. Additionally, distinct perturbations in the eCB system have been reported among men and women undergoing cannabis withdrawal^15^.

Similarly, several studies have reported sex-differences in the levels of peripheral inflammatory markers among individuals with PTSD^16,17^. A pro-inflammatory state appears to characterize PTSD, as supported by an increased prevalence of autoimmune disorders among affected individuals^18^, a heightened expression of pro-inflammatory genes^19–21^, and higher levels of cytokines compared to controls [for a review see Michopoulos 2017^22^]. Among the few studies exploring sex differences in inflammatory markers in individuals with PTSD, Spitzer et al. (2010) and Miller et al. (2018) both reported higher levels of C-reactive protein (CRP) in women compared to men. However, findings on other inflammatory markers, such as interleukin-6 (IL-6) and interleukin-10 (IL-10), have been inconsistent, showing either no differences among sexes^23^ or lower levels in women than men in the aftermath of traumatic stress exposure^24^.

Despite preliminary evidence of sex-differences in the eCB system and in the inflammatory response, it remains unclear whether PTSD is linked to distinct biological signatures in men and women. To begin addressing these knowledge gaps, our study examined circulating eCB levels and peripheral pro-inflammatory markers in a cohort of men and women with PTSD compared to non-psychiatric controls. Analyses were conducted in the entire sample and in the subgroup of individuals with comorbid major depressive disorder (MDD), given high comorbidity rates between MDD and PTSD and evidence of perturbations in eCB system and heightened inflammation in subjects affected by this condition. We hypothesized that women with PTSD, with and without comorbid MDD, would exhibit increased levels of pro-inflammatory molecules compared to both men with PTSD and healthy controls. Further, we hypothesized that men with PTSD, with and without comorbid MDD, would have lower concentrations of eCBs compared to male healthy controls, while no differences in eCB levels would be observed in women with PTSD compared to both men with PTSD and female healthy controls.

## 2. Material and Methods

### 2.1 Subject and sample selection

We conducted a retrospective cohort study using demographic, clinical and genotype data, and serum samples, from the Mass General Brigham (MGB) Biobank, a large, ongoing biorepository that recruits patients from hospitals and clinics across the MGB system. Eligible participants are adult patients who provide informed consent by signing a Data and Sample Use Agreement, allowing researchers to access their de-identified health records. The MGB Biobank includes biospecimens linked to extensive electronic health record (EHR) and survey data from over 135,000 participants and has been widely utilized in diverse research studies^25^. The quality of EHR data is validated through quality assurance measures and machine learning-derived phenotypes^26^. This study was approved by the Mass General Brigham Institutional Review Board.

Using the MGB Biobank Portal Query tool, we compiled a cohort of 176 individuals aged 18–45 prospectively selected to achieve a sex ratio of approximately 1:1. Of those, 88 subjects were healthy controls (“healthy controls” or HC) and 88 received a diagnosis of PTSD at the Brigham and Women’s Hospital and/or at the Massachusetts General Hospital between January 2015 and December 2021 (“PTSD patients”). For all subjects, EHR data and serum samples were available. PTSD diagnosis was determined by the presence of at least two entries of ICD-10 (International Classification of Diseases, 10th edition) code in a patient’s HER and by a medication history consistent with such diagnosis. Exclusion criteria for both the PTSD and HC groups included history of autoimmune, endocrine and/or chronic inflammatory disorders; diagnosis of bipolar disorder, schizophrenia and other psychotic disorders, as assessed by the respective ICD-10 codes; use of medications known to alter inflammatory response and/or eCB levels at the time the biospecimens were collected; body mass index > 30.

### 2.2 Measures

#### Endocannabinoids

Serum samples underwent stable isotope dilution liquid chromatography/tandem mass spectrometry (LC-MS/MS) to assess circulating levels of eCBs, specifically Anandamide (AEA), 2-Arachidonyl glycerol (2-AG), Oleoylethanolamide (OEA), and Arachidonic acid (AA). Briefly, 100 µL serum was incubated at −20°C for 10 min with 900 ml ice-cold acetone and 400 μL Tris buffer (50 mM, pH 8.0) to precipitate proteins. After spinning at 2000g at 4°C for 10 min, the supernatant was transferred to a glass tube to evaporate the acetone phase under nitrogen flow. Then, 0.5 mL of ice-cold methanol/Tris buffer (50 mM, pH 8.0) added to the supernatant, 1:1, containing 7 ng of [2H4] arachidonoyl ethanolamide ([2H4] AEA), and 50 ng of [2H5] arachidonoyl glycerol ([2H5] 2AG), as internal standards. Then, the supernatant was extracted two times with 2 ml of CHCl3:MeOH (2:1, vol/vol). Lower chloroform phase was collected and transfer another glass tube. Then combined chloroform phases were dried under nitrogen flow. The dried samples were reconstituted in 50 μL of ice-cold methanol prior to loading autosampler for mass spectrometry measurements.

LC-MS/MS analyses were conducted on an Agilent 6470 triple quadrupole mass spectrometer (Agilent Technologies) coupled to an Agilent 1200 LC system. Liquid chromatographic separation was obtained using 2 μL injections of samples onto a InfinityLab Poroshell 120 EC-C18 column (3.0mm×100 mm, 2.7 Micron) from the Agilent Technologies. The autosampler temperature was set at 4◦C and the column was maintained at 34◦C during the analysis. Gradient elution mobile phases consisted of 0.1% formic acid in H2O (phase A) and 0.1% formic acid in MeOH (phase B). Gradient elution (350 μL/min) was initiated at 10% B, followed by a linear increase to 50% B at 0.5 min, followed by a linear increase to 85% B at 3 min and maintained until 14 min, then increased linearly to 100% B at 18 min and maintained until 20 min. The mass spectrometer was set for electrospray ionization operated in positive ion mode. The source parameters were as follows: capillary voltage, 3,500 V; gas temperature, 300 °C; drying gas, 5 L/min; nitrogen was used as the nebulizing gas. Collision-induced dissociation was performed using nitrogen. Levels of each compound were analyzed by multiple reaction monitoring. The molecular ion and fragment for each compound were measured as follows: m/z 348.3→62.1 for AEA, m/z 379.3→287.2 for 2AG, m/z 384.3→91.1 for [^2^H_5_] 2-AG, m/z 352.3→66.1 for [^2^H_4_] AEA, m/z 305.2➔91.1 for AA, and m/z 326.3➔62.1 for OEA. The analytes were quantified using MassHunter Workstation LC/QQQ Acquisition and MassHunter Workstation Quantitative Analysis software (Agilent Technologies). Levels of analytes in the samples were measured against standard curves and were expressed as pmol/ml. One participant was missing endocannabinoid values; these missing values were ignored for statistical tests.

#### Inflammatory Markers

Serum concentration of several pro-inflammatory molecules (IL-1β, IL-6, IL-8, IL-18, TNF-α and CRP) was quantified using the validated Meso Scale Discovery Platform (Rockville, MD, USA) assays by the Laboratory of Genital Tract Biology at Brigham & Women’s Hospital, under accreditation by the College of American Pathologists, as previously described^27^. All samples were above the assay’s lower limit of detection for each analyte. Analyte concentrations were expressed as mg/L for CRP and pg/mL for IL-1β, IL-6, IL-8, IL-18, and TNF-α. Two participants were missing inflammatory marker values; thus, they were excluded from further analyses.

#### C385A Carrier Status

The C385A single nucleotide polymorphism (SNP) is relatively common, and the presence of the recessive A allele has been associated with reduced catabolic efficiency of the FAAH enzyme, leading to elevated anandamide (AEA) levels^28,29^. To explore this, we obtained genotype data, where available, to assess the frequency of the C385A allele by sex and disease status. Genotyping was conducted by the biobank using Illumina’s multi-ethnic genotyping arrays, including the MEGA array, MEGA Ex array, and MEG BeadChip array. Genotype data were available for a subset of the sample (71 of 119 PTSD patients and 48 of 119 healthy controls), with ∼31% of the sample missing genotype information.

Due to the low frequency of individuals homozygous for the A allele, genotypes were collapsed into two groups for analysis: C385 homozygotes (CC) and carriers of the 385A allele (AX).

#### Other variables

Smoking status, comorbid diagnosis of cannabis use disorder, alcohol use disorder and major depressive disorder were extracted from EHRs using the respective ICD-10 codes.

### 2.3 Statistical Analyses

To characterize the study population, we examined differences in demographics and clinical characteristics between PTSD patients and controls, overall and by sex, using Fisher’s exact test for binary variables or Student’s t-test and the Wilcoxon rank-sum test for continuous variables. Smoking status and psychiatric comorbidities were coded as binary variables (e.g., 0=non-smoking, 1=smoking). Additionally, body-mass index (BMI) was recorded as a continuous variable with responses ranging from 19.5 to 46.7. Biological sex was collected as a binary measure (female/male), whereas race and ethnicity were collected as categorical variables which were collapsed into four and three categories, respectively.

Distribution of peripheral inflammatory and endocannabinoid analytes were descriptively examined, including assessing correlations between analyte levels within sex and PTSD/HC groups (Table S3). Analyte levels were log2 transformed to assess proportional change in these markers. Linear regression was then used to predict differential abundance of each analyte between PTSD patients and HCs within each sex (Δ *females* and Δ *males)*, adjusting for age. Two-sided t-tests were conducted, with the Benjamini-Hochberg procedure applied to adjust for multiple comparisons. Given that analytes in the eCB and inflammatory groups are biologically connected, we applied the FRY method, a self-contained gene set test that does not assume that genes are expressed independently of one another^30^, using the Limma package in *R*^31^. This method enabled us to assess the differential abundance of endocannabinoids and inflammatory markers collectively, rather than examining each analyte separately. The level of statistical significance was set at p<0.05 for all tests. Descriptive statistical analyses were conducted using Stata/SE 18 (StataCorp, College Station, TX), while all other analyses were performed using R version 4.3.1 (2023).

## 3. Results

### 3.1 Sample characteristics

The final cohort included 173 individuals: 88 PTSD patients and 85 HC (Table S1). This sample was selected to have a male-to-female ratio close to 1:1 and therefore consists of 90 males (52%; 46 with PTSD) and 83 females (48%; 42 with PTSD). The majority of individuals were non-Hispanic white (69.9%) and nonwhite (11%) with a median age of 31 years old. The distributions of these demographics did not significantly differ between PTSD patients and HCs. Among participants with available BMI data (9/34 controls, 25/34 patients), the median BMI was 25.1 kg/m^2^ with no significant differences observed between the PTSD and HC groups. As expected, the PTSD group had significantly higher rates of smoking (*p<0.001*), cannabis use disorder (*p<0.05*), alcohol use disorder (*p<0.001*), and comorbid major depressive disorder (MDD) (*p<0.001*) compared to HCs. Among psychiatric comorbidities, MDD was the most prevalent (54.8% females with PTSD, 56.5% males with PTSD).

Regarding *FAAH* C385A carrier status, 41 individuals (34.5%) were either heterozygous (AC) or homozygous recessive (AA) and 54 individuals (31.2%) did not have genotype data available. Among individuals with available data, the genotype distribution deviated significantly from Hardy Weinberg Equilibrium (χ² = 5.79, *p* < 0.05). *FAAH* 385A allele frequencies did not differ between the PTSD and HC groups. Next, we examined sex-differences in demographic and baseline characteristics within each group (PTSD and HC) (Table S2). No significant differences were found, with the exception of age. Indeed, male patients were slightly older than female patients (*p<0.01*). Similarly, male controls were older that female controls (*p <0.01*).

Lastly, when comparing PTSD and HC groups across sexes, we observed that both male and female individuals with PTSD had significantly higher rates of smoking (*p <0.001*) and comorbid MDD (*p* < 0.001) compared to their respective HC groups. Furthermore, males with PTSD exhibited significantly higher rates of alcohol use (*p < 0.001*) compared to male HCs.

**Table 1.** Sample Characteristics by Sex and Disease State.

|  | Females |  |  |  | Males |  |  |  |
| --- | --- | --- | --- | --- | --- | --- | --- | --- |
|  | Total | HC | PTSD | HC vs PTSD | Total | HC | PTSD | HC vs PTSD |
|  | N=83 | N=41 | N=42 |  | N=90 | N=44 | N=46 |  |
|  | *N (%) or Median (IQR)* |  |  | *signif.* | *N (%) or Median (IQR)* |  |  | *signif.* |
| Race |  |  |  |  |  |  |  |  |
| Non-Hispanic white | 54 (65.1%) | 27 (65.9%) | 27 (64.3%) |  | 67 (74.4%) | 31 (70.5%) | 36 (78.3%) |  |
| Nonwhite | 9 (10.8%) | 5 (12.2%) | 4 (9.5%) |  | 10 (11.1%) | 8 (18.2%) | 2 (4.3%) |  |
| Other | 4 (4.8%) | 1 (2.4%) | 3 (7.1%) |  | 2 (2.2%) | 0 (0.0%) | 2 (4.3%) |  |
| Unknown• | 16 (19.3%) | 8 (19.5%) | 8 (19.0%) |  | 11 (12.2%) | 5 (11.4%) | 6 (13.0%) |  |
| Ethnicity |  |  |  |  |  |  |  |  |
| Non-Hispanic | 58 (69.9%) | 30 (73.2%) | 28 (66.7%) |  | 65 (72.2%) | 33 (75.0%) | 32 (69.6%) |  |
| Hispanic | 3 (3.6%) | 1 (2.4%) | 2 (4.8%) |  | 3 (3.3%) | 3 (6.8%) | 0 (0.0%) |  |
| Unknown• | 22 (26.5%) | 10 (24.4%) | 12 (28.6%) |  | 22 (24.4%) | 8 (18.2%) | 14 (30.4%) |  |
| Age (yr.) | 28.0 (25.0-33.0) | 28.0 (25.0-33.0) | 28.0 (26.0-32.0) |  | 34.0 (28.0-43.0) | 35.0 (28.0-43.0) | 33.0 (28.0-42.0) |  |
| BMI (kg/m<sup>2</sup>) | 23.2 (20.9-28.8) | 23.2 (19.5-24.7) | 23.3 (20.9-29.5) |  | 26.8 (23.6-28.5) | 25.8 (23.2-28.2) | 26.8 (24.3-28.7) |  |
| Smoking | 22 (26.5%) | 0 (0.0%) | 22 (52.4%) | *** | 14 (15.6%) | 0 (0.0%) | 14 (30.4%) | *** |
| Cannabis | 2 (2.4%) | 0 (0.0%) | 2 (4.8%) |  | 4 (4.4%) | 0 (0.0%) | 4 (8.7%) |  |
| Alcohol | 5 (6.0%) | 0 (0.0%) | 5 (11.9%) |  | 13 (14.4%) | 0 (0.0%) | 13 (28.3%) | *** |
| Depression | 23 (27.7%) | 0 (0.0%) | 23 (54.8%) | *** | 26 (28.9%) | 0 (0.0%) | 26 (56.5%) | *** |
| C385A Carriers† |  |  |  |  |  |  |  |  |
| C-C | 38 (70.4%) | 16 (72.7%) | 22 (68.8%) |  | 40 (61.5%) | 12 (46.2%) | 28 (71.8%) |  |
| A-X | 16 (29.6%) | 6 (27.3%) | 10 (31.2%) |  | 25 (38.5%) | 14 (53.8%) | 11 (28.2%) |  |

\*\*\* $p < 0.001$ , \*\* $p < 0.01$ , \* $p < 0.05$
•Unknown category includes declined and missing/unavailable responses.
†AX: AC or AA; N= 54 females (22 healthy controls [HC], 32 patients); 65 males (26 HCs, 39 PTSD)

### 3.2 Differential Abundance of Inflammatory Markers and Endocannabinoid Levels

#### Male PTSD vs controls (Δ males)

Among males, concentrations of all inflammatory markers except TNFα (*q<0.05*) did not differ between patients with PTSD and controls (Table 3). In contrast, endocannabinoid levels were significantly reduced in patients, as compared to controls, including AA (*q<0.01*), AEA (*q<0.001*), and OEA (*q<0.001*), though no significant differences was observed for 2-AG. Full results, including nominal p-values and log2 fold change, are presented in Table S4. Since *FAAH* genotype influences circulating levels of AEA, we conducted additional regression analyses controlling for this variable (Table S6). These analyses, restricted to the subset of individuals with available genotype data, yielded results consistent with the primary models.

**Table 3.** Linear regression models predicting change in endocannabinoid and inflammatory markers between PTSD patients and healthy controls by sex.

| N | Females |  |  |  |  | Males |  |  |  |  |
| --- | --- | --- | --- | --- | --- | --- | --- | --- | --- | --- |
|  | HC | PTSD | Δ Females |  |  | HC | PTSD | Δ Males |  |  |
|  | 41 | 42 | 83 |  |  | 44 | 46 | 90 |  |  |
|  | Avg. - log2 scale |  | FC | q-value | signif. | Avg. - log2 scale |  | FC | q-value | signif. |
| Endocannabinoids |  |  |  |  |  |  |  |  |  |  |
| AA | 9.60 | 9.90 | 1.239 | 0.173 |  | 10.10 | 9.56 | -1.448 | 0.008 | ** |
| AEA | 0.41 | 0.53 | 1.088 | 0.429 |  | 0.93 | 0.47 | -1.370 | 0.000 | *** |
| OEA | 2.55 | 2.48 | -1.053 | 0.719 |  | 2.83 | 2.23 | -1.501 | 0.000 | *** |
| 2-AG | 3.91 | 4.44 | 1.452 | 0.025 | * | 4.84 | 4.80 | -1.017 | 0.898 |  |
| Inflammatory Markers |  |  |  |  |  |  |  |  |  |  |
| CRP | 0.92 | 0.87 | -1.026 | 0.937 |  | 0.46 | 0.63 | 1.181 | 0.744 |  |
| IL1β | -2.66 | -2.41 | 1.191 | 0.386 |  | -2.45 | -2.61 | -1.116 | 0.623 |  |
| IL6 | 0.25 | 1.07 | 1.776 | 0.013 | * | 0.66 | 0.84 | 1.160 | 0.623 |  |
| IL8 | 3.23 | 3.96 | 1.666 | 0.025 | * | 3.23 | 3.62 | 1.318 | 0.260 |  |
| IL18 | 8.99 | 9.01 | 1.018 | 0.937 |  | 9.22 | 9.23 | 1.018 | 0.898 |  |
| TNFα | -0.39 | 0.24 | 1.544 | 0.029 | * | -0.17 | 0.44 | 1.515 | 0.031 | * |

\*\*\* $p < 0.001$ , \*\* $p < 0.01$ , \* $p < 0.05$
HC: healthy controls; FC: fold change

#### Female PTSD vs controls (Δ females)

Among females, concentrations of IL-6 (*q<0.05*), IL-8 (*q<0.05*), and TNFα (*q<0.05*) significantly higher in PTSD patients compared to HCs after controlling for age (Table 3). In contrast, levels of CRP, IL-1β, and IL-18 did not significantly differ between the PTSD and HC groups. With respect to endocannabinoids, no significant differences were found for AA, AEA, and OEA. However, 2-AG was significantly elevated in female PTSD patients compared to female HCs (*q<0.05*). Similar to the male subgroup, additional regression models controlling for *FAAH* genotype in females revealed largely consistent results, as presented in Table S6.

#### Subgroup analyses: comorbid PTSD and MDD

Next, regression models were applied to a subset of participants with concurrent diagnoses of both PTSD and MDD (Table 4, full results in Table S5). In this smaller sample, the results largely mirrored those observed in the broader cohort. Specifically, IL-8 (q < 0.05) was predicted to be elevated in female PTSD patients compared to female controls, while AEA (q < 0.05) and OEA (q < 0.05) were predicted to be significantly depleted in male PTSD patients compared to male controls. Due to the reduced sample size, further stratification and modeling by *FAAH* genotype were not performed.

**Table 4.** Linear regression models predicting change in endocannabinoid and inflammatory markers among individuals with comorbid MDD and PTSD as compared to healthy controls.

| N | $\Delta$ Females<br>23 | | | $\Delta$ Males<br>26 | | |
| --- | --- | --- | --- | --- | --- | --- |
|  | *FC* | *q-value* | *signif.* | *FC* | *q-value* | *signif.* |
| Endocannabinoids |  |  |  |  |  |  |
| AA | 1.356 | 0.143 |  | -1.348 | 0.086 |  |
| AEA | 1.013 | 0.886 |  | -1.315 | 0.011 | * |
| OEA | -1.043 | 0.844 |  | -1.378 | 0.011 | * |
| 2-AG | 1.253 | 0.338 |  | 1.115 | 0.594 |  |
| Inflammatory Markers |  |  |  |  |  |  |
| CRP | -1.605 | 0.338 |  | 1.373 | 0.594 |  |
| IL1β | 1.214 | 0.338 | | -1.001 | 0.992 | |
| IL6 | 1.502 | 0.143 |  | 1.169 | 0.594 |  |
| IL8 | 1.856 | 0.039 | * | 1.056 | 0.872 |  |
| IL18 | -1.037 | 0.844 |  | 1.120 | 0.594 |  |
| TNFα | 1.272 | 0.338 | | 1.143 | 0.594 | |

\*\*\* $p < 0.001$ , \*\* $p < 0.01$ , \* $p < 0.05$
HC: healthy controls; FC: fold change

### 3.3 Group-level analysis results

Considering that both individual eCB and inflammatory markers are functionally related and that, therefore, they exhibit some degree of correlation, we next analyzed sex-differences between patients and controls across the entire group of eCBs and inflammatory molecules, respectively. Consistent with the regression results for individual analytes, female PTSD patients did not show significant differences in eCB levels compared to female HCs. However, male PTSD patients exhibited significantly lower eCB levels compared to male HCs (*p* < 0.001). This pattern was also observed in individuals with comorbid MDD, although only marginally significant (*p* < 0.05) (Table 5).

**Table 5.** Differential abundance of eCB and inflammatory markers among all individuals and those with comorbid MDD.

|  | Overall Sample<br>N=173 |  |  | Comorbid MDD<br>N=49 |  |  |
| --- | --- | --- | --- | --- | --- | --- |
|  | *Prop. of Diff. Markers†* | *q-value* | *signif.* | *Prop. of Diff. Markers†* | *q-value* | *signif.* |
| **Endocannabinoids** |  |  |  |  |  |  |
| Δ Females | 0.25 | 0.104 |  | 0.25 | 0.261 |  |
| Δ Males | - 0.75 | 0.000 | *** | - 0.75 | 0.024 | * |
| **Inflammatory Markers** |  |  |  |  |  |  |
| Δ Females | + 0.50 | 0.028 | * | 0.33 | 0.261 |  |
| Δ Males | 0.17 | 0.211 |  | 0 | 0.350 |  |

\*\*\* $p < 0.001$ , \*\* $p < 0.01$ , \* $p < 0.05$
†proportion of markers with significantly different concentrations in PTSD vs HC ( $p < 0.05$ )
Endocannabinoids: AA, AEA, OEA, 2-AG
Inflammatory markers: IL-1β , IL-6, IL-8, IL-18, TNFα , CRP

For the inflammatory molecules group, female PTSD patients demonstrated significantly higher levels of such markers (*p*< 0.05) compared to female HCs. In contrast, no significant differences were observed between male PTSD patients and male HCs. Among individuals with comorbid depression, no significant differences were found in either sex.

### 3.4 Sex Differences in Levels of Endocannabinoid and Inflammatory Markers

Lastly, we compared the magnitudes of group-differences (PTSD *vs* HC) in analyte levels observed within the female and male subsamples (*Δ females - Δ males*) (Table 6, group averages presented in Figures 1 and 2). We identified significant sex differences in aggregate eCB levels (*p* <0.001), driven by differential changes in AA (*p* <0.01), AEA (*p* <0.01), and OEA (*p* <0.05), but not 2-AG (*p* =0.1). Conversely, the magnitude of group-differences in inflammatory markers did not significantly differ between the female and male subsamples (*p’s>0.05*) [Table 6]. Similar results were obtained when analyses were performed including only individuals with comorbid MDD.

**Table 6.** Sex differences in endocannabinoid and inflammatory marker concentrations between the female and male subgroups (PTSD *vs* HC; *Δ females - Δ males*)

|  | Overall Sample<br>N=173 |  | Comorbid MDD<br>N=49 |  |
| --- | --- | --- | --- | --- |
|  | *q-value* | *signif.* | *q-value* | *signif.* |
| Endocannabinoid Group | 0.000 | *** | 0.022 | * |
| AA | 0.004 | ** | 0.021 | * |
| AEA | 0.004 | ** | 0.127 |  |
| OEA | 0.021 | * | 0.149 |  |
| 2-AG | 0.105 |  | 0.653 |  |
| Inflammatory Marker Group | 0.350 |  | 0.672 |  |
| CRP | 0.838 |  | 0.229 |  |
| IL1β | 0.269 | | 0.483 | |
| IL6 | 0.163 |  | 0.483 |  |
| IL8 | 0.531 |  | 0.149 |  |
| IL18 | 0.998 |  | 0.483 |  |
| TNFα | 0.998 | | 0.693 | |

\*\*\* $p < 0.001$ , \*\* $p < 0.01$ , \* $p < 0.05$
FC: fold change

**Figure 1.**
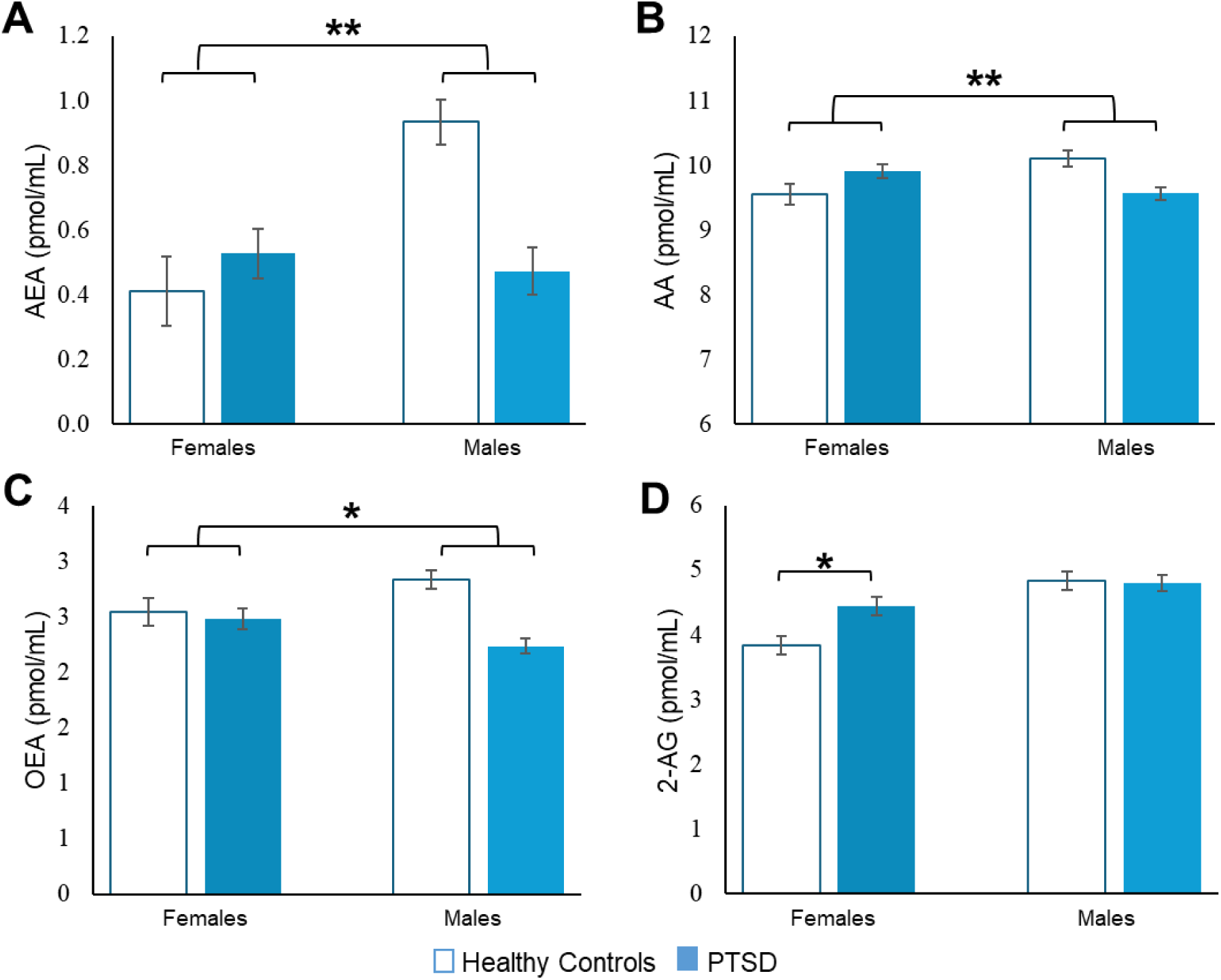
Sex differences in circulating endocannabinoids characterized by decreased AEA (A; p=0.004), AA (B; p=0.004), and OEA (C; p=0.021) in males with PTSD. Females with PTSD showed increased 2-AG, compared to female HC (D, p=0.025). Data represent log2-transformed means ± SEM in pmol/mL.

**Figure 2.**
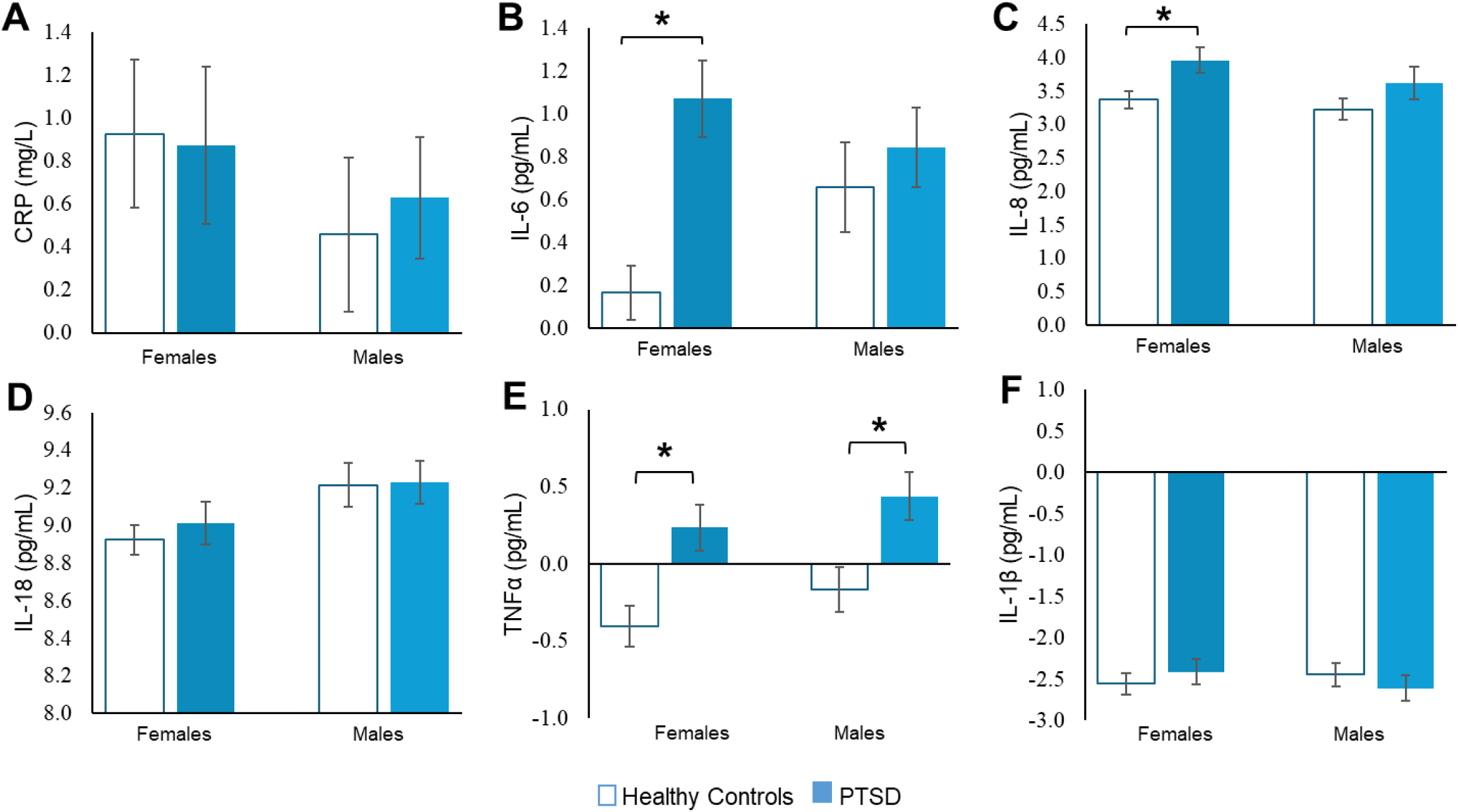
Higher concentrations of circulating IL-6 (B, p=0.013), IL-8 (C, p=0.025), and TNFα (E, p=0.029) in females with PTSD, compared to female HC. Greater concentration of TNFα (E, p=0.031) in males with PTSD, compared to male HC. Data represent log2-transformed means ± SEM in pg/mL or mg/L.

## 4. Discussion

To our knowledge, this is the first study to examine circulating eCB levels and peripheral pro-inflammatory markers in men and women with PTSD compared to non-psychiatric controls, as well as sex-differences in these measures. Consistent with our hypothesis, reduced eCB levels-- particularly anandamide (AEA) and its metabolites oleoylethanolamide (OEA) and arachidonic acid (AA) - were associated with PTSD diagnosis exclusively in men, while no differences in eCB levels were observed between female patients and controls or between male and female patients with PTSD. In contrast, female patients showed elevated levels of several pro-inflammatory markers (IL-6, IL-8, and TNF-α) compared to female controls, whereas male patients exhibited only a modest increase in TNF-α levels compared to male controls. The magnitude of the group differences (PTSD *vs* HC) did not significantly vary between sexes. Additionally, male and female patients with comorbid PTSD and MDD exhibited similar alterations in levels of eCBs and inflammatory markers compared to controls, suggesting shared biological mechanisms underlying these conditions.

Our finding that PTSD is characterized by a depletion in eCB levels is in line with numerous preclinical and clinical studies showing that exposure to chronic stress reliably reduces central and peripherical levels of AEA ^32^. With respect to functionality, lower levels of circulating AEA have been found to correlate with higher anxiety ^33,34^ and with greater symptom severity in individuals with PTSD ^10^. Research examining the effects of elevated AEA in humans, through either pharmacological or genetic means [i.e., the *FAAH* C385A SNP that results in higher levels of AEA ^29^], has shown that reduced activation of the amygdala in response to threat cues, reduced anxiety following exposure to acute stress, decrease in PTSD hyperarousal symptoms, increased extinction of fear memories and improved fear extinction recall are all associated to an increase in AEA levels ^11,12,35–37^.

Interestingly, a recent community-based cohort study, predominantly enrolling male individuals (∼70%), reported that serum anandamide (AEA) concentrations at the time of traumatic injury were significantly higher in those who developed PTSD at follow-up (6-8 months later)^38^. Conversely, a study in women found no differences in eCB levels between those with PTSD and healthy controls^39^. These findings align with our data showing that male controls exhibit significantly higher AEA levels compared to both male PTSD patients and the female subsamples (PTSD and HCs). This suggests that AEA tone is physiologically elevated in males, as also supported by evidence from a large-scale study demonstrating higher AEA levels in males compared to females across the lifespan^14^. Thus, eCB deficits in response to chronic stress and trauma exposure may represent a male-specific mechanism underlying PTSD, whereas AEA signaling may not play a central role in fear and stress response in females. Notably, a study found that augmenting AEA signaling in female rodents impairs fear extinction^13^. Future studies should examine whether sex differences in AEA levels contribute to dysregulated fear learning and heightened anxiety in a sex-specific manner. Additionally, it will be important to investigate the role of other eCBs, specifically 2-AG, the other primary eCB ligand, in modulating fear and stress processes in females and males.

We further observed that both female and male patients with PTSD exhibited increased levels of pro-inflammatory markers compared to their control groups. These findings parallel previous reports indicating that individuals with PTSD exhibit elevated levels of pro-inflammatory molecules, including IL-1β ^40–42^, IL-2 ^43^, IL-6^44–46^ and TNF-α ^44^. However, some studies have found no clear link between PTSD and heightened inflammation (for a review, see Michopoulos et al., 2017). A meta-analysis of 20 studies comparing individuals with PTSD and healthy controls reported significantly higher levels of IL-1β, IL-6, TNF-α, and IFN-γ in those with PTSD, regardless of comorbid major depression^9^, consistent with our findings, thus providing robust evidence of an association between PTSD and heightened inflammation.

However, despite it is well-established that immune responses and inflammation strongly differ between sexes^47^, the influence of biological sex in modulating inflammatory response in PTSD has been largely overlooked. Two studies reported higher CRP circulating levels in women with PTSD *vs* men with PTSD^16,17^. Moieni and colleagues (2015) reported that endotoxin-induced increases in IL-6 and TNF-*α* levels correlated with greater depressed mood and social disconnection only in women^48^. A further study examining changes in immune cell gene expression found no evidence for chronic inflammation in male PTSD subjects, while women with PTSD exhibited a gene expression pattern suggestive of increased activity of pathways related to immune activation^49^. In contrast, no differences in circulating levels of IL-6 and IL-10 were observed in 33 men *vs* 8 women with PTSD^23^. While several of these studies were not designed and/or powered to examine sex differences, preclinical data suggest that psychological trauma-induced inflammation is more robust in female vs male rodents [for a review see Levesque et al., 2023^50^]. In our cohort, female individuals with PTSD showed a robust increase in several pro-inflammatory markers (i.e., IL-6, IL-8, and TNF-α), while within the male subsample, only TNF-α were significantly higher in PTSD patients *vs* HCs. However, no sex differences were detected when comparing the magnitudes of group differences in these markers between the female and male subsamples. This seemingly inconsistent finding may be attributed to several factors. First, PTSD has been linked to a state of low-grade inflammation^22^, which could result in moderate differences in pro-inflammatory markers between patients and controls, particularly when assessed at rest. Notably, recent evidence indicates that levels of inflammatory molecules, such as IL-6, are directly influenced by stress exposure and during fear extinction processes^50^. Therefore, assessing the magnitude of sex differences in immune responses to trauma may require experimental stress or fear-provoking paradigms able to provoke an inflammatory response.

Lastly, we also explored whether comorbidity between MDD and PTSD was associated with similar alterations in eCB and inflammatory markers as those observed in individuals with PTSD alone. A prior meta-analysis evaluating sex differences in inflammatory markers in PTSD^9^, considered comorbid MDD as a potential confounder. However, MDD and PTSD share numerous features, including dysregulated eCB signaling^51^, heightened inflammation^8^, and higher prevalence in women^52^. These shared characteristics suggest overlapping neurobiological mechanisms, a hypothesis supported by our results that indicate similar sex-specific alterations in levels of eCBs and inflammatory markers in male and female individuals with PTSD with and without comorbid MDD.

Our findings should be interpreted in light of several limitations. First, while a relative large sample size is a strength of this study, diagnostic and clinical data were obtained from electronic medical records and not from prospectively delivered structured clinical interviews and exams. We, therefore, cannot determine with certainty the reliability of all diagnoses (or absence thereof). However, our cohort was sampled from two hospitals offering specialist psychiatric services. Furthermore, we selected only individuals who had at least two ICD-10 entries for PTSD in their EHRs and we used subjects ‘medication history as a proxy to confirm diagnosis of PTSD, thus we expect misdiagnosis rates to be in the lowest range. Second, we did not have access to measures of symptom severity, as well as data regarding exposure to childhood trauma, which should be considered in future research. These limitations are inherent to retrospective cohort studies using EHRs, which, however, offer an extremely valuable opportunity to explore novel research questions and could help inform prospective studies. It should also be noted that we were unable to assess the relationship between gonadal sex hormone levels and eCBs or inflammatory markers, despite evidence suggesting that sex hormones modulate levels of both markers^53,54^. Although we restricted our analyses to individuals aged 18-45 years to account for the influence of hormones and age, future studies should investigate the extent by which sex hormones contribute to the observed sex -specific alterations in eCBs and inflammatory molecules.

In summary, our study is the first to demonstrate that men and women with PTSD exhibit distinct alterations in endocannabinoids, particularly AEA, and inflammatory markers, suggesting sex-specific contributions to PTSD pathophysiology. Given that PTSD is among the most sex-polarized psychiatric disorders^1^, yet the biological basis of this difference remains poorly understood, future research should build on these findings to explore whether distinct biological perturbations underlie dysregulated fear and stress processes.

## Data Availability

All data produced in the present study are available upon reasonable request to the authors.

## Acknowledgements

This work has been supported by the Mary Ann Tynan Fellowship Program and the Women’s Brain Initiative/Brigham and Women’s Hospital. The authors acknowledge the participants and administrators of the Mass General Brigham Biobank for their contribution to this work. The authors would also like to thank Enterprise Research Infrastructure & Services at Mass General Brigham for the provision of analysis software, Robert James Glynn, PhD, ScD for valuable feedback, and support from UM1TR004408 award through Harvard Catalyst Biostatistical Consulting Program and financial contributions from Harvard University and its affiliated academic healthcare centers. The content is solely the responsibility of the authors and does not necessarily represent the official views of Harvard Catalyst, Harvard University and its affiliated academic healthcare centers, or the National Institutes of Health.

